# An experimental feasibility study evaluating the adequacy of sportswear-type wearables for recording physical exercise intensity

**DOI:** 10.1101/2021.12.27.21268299

**Authors:** Yoshihiro Marutani, Shoji Konda, Issei Ogasawara, Keita Yamasaki, Teruki Yokoyama, Etsuko Maeshima, Ken Nakata

**Affiliations:** Graduate School of Sport Science, Osaka University of Health and Sport Sciences, Kumatori, Osaka, Japan, 590-0496; Department of Health and Sport Sciences, Osaka University Graduate School of Medicine, Toyonaka, Osaka, Japan, 560-0043; Department of Sports Medical Biomechanics, Osaka University Graduate School of Medicine, Suita, Osaka, Japan, 565-0871

**Keywords:** wearable sensor, acceleration, electrocardiogram, heart rate, exercise intensity

## Abstract

Sportswear-type wearables with integrated inertial sensors and electrocardiogram (ECG) electrodes, have been developed. We examined the feasibility of using sportswear-type wearables to evaluate exercise intensity within a controlled laboratory setting. Six male college athletes were asked to wear a sportswear-type wearable while performing a treadmill test that reached up to 20 km/h. The magnitude of the filtered tri-axial acceleration signal, recorded by the inertial sensor, was used to calculate the acceleration index. The R-R intervals of ECG were used to determine heart rate; the external validity of the heart rate was then evaluated according to oxygen uptake, which is the gold standard physiological exercise intensity. Single regression analyses between treadmill speed and the acceleration index in each participant showed that the slope of the regression line was significantly greater than zero with a high coefficient of determination (walking, 0.95; jogging, 0.96; running, 0.90). Another single regression analyses between heart rate and oxygen uptake showed that the slope of the regression line was significantly greater than zero, with a high coefficient of determination (0.96). Together, these results indicate that sportswear-type wearables are a feasible technology for evaluating physical and physiological exercise intensity across a wide range of physical activities and sport performances.

## 1. Introduction

The benefits of wearable sensors have attracted attention worldwide. With the ability to monitor the intensity of physical activity in real time, the field of sports medicine has recognized that wearables are technically superior to conventional monitoring techniques, such as subjective questionnaires [1-4]. Wearables thus allow exercise management and training programs to be quantitatively analyzed [5-7]. Hence, most wearables today use a combination of physical and physiological indices. The former evaluates physical intensity, determined using a global positioning system (GPS) and accelerometer to quantify displacement, speed, and acceleration [8-10]. Conversely, the percentage of maximum heart rate (%HRmax), and heart rate reserve (HRR) represent physiological intensity [11, 12]. Together, these indices allow exercise intensity to be measured. Further, the effects of exercise have traditionally been measured via oxygen uptake [13]; however, this monitoring method is not suitable for non-experimental settings. As a result, alongside the rise in the use of wearables, heart rate indices – which allow non-invasive, day-to-day observations – have become the preferred monitoring method over oxygen uptake [14-16]. Therefore, in recent years, the heart rate indices have been widely used in daily life as a non-invasive method in the field of wearable sensing [14-16].

The most popular consumer wearable is the watch-type sensor worn on the wrist, which allows the physical and physiological indices of daily living, exercise, and sports activity to be recorded [17, 18]. The second most common is the chest-belt-type wearable, which has been used for research purposes and to monitor exercise intensity in competitive sports [19, 20]. Currently, athletic associations – such as the International Federation of Association Football (FIFA) and International Tennis Federation (ITF) – have introduced rules allowing players to don wearables during official games. The downside, however, is that both watch- and chest-belt-type sensors cause discomfort and increase the risk of injury, for example in situations where players come into contact with each other. The development of wearables with improved comfort and lower risk of injury will enable the quantification of exercise intensity across a wide range of physical activities, from daily life to competitive sports [21, 22].

Recently, a sportswear-type wearable with a small inertial sensor attached to the chest and two ECG electrodes sewn on opposite sides, has been developed [23]. A major advantage of sportswear-type wearables is that it is as comfortable as sportswear, which enables athletes to wear them in daily training, practice and even in competitive match. A previous study had demonstrated the feasibility of recording ECG signals while running [24, 25]; however, the feasibility of evaluating exercise intensity using an embedded inertial sensor is not yet known. With our study, we aimed to examine this across a wide range of physical activities, within a controlled laboratory setting.

## 2. Methods

### 2.1 Participants

Six male college athletes (mean age ± SD: 18.8 ± 0.4 years; mean height ± SD: 171.2 ± 7.1 cm; mean weight ± SD: 62.2 ± 5.5 kg; mean BMI ± SD: 19.1 ± 4.0 kg/m^2^) volunteered to participate in this study. Participants included were: (1) collegiate male athletes who exercised regularly; (2) were disease-free and in good health; (3) previous experience with a treadmill; (4) ability to complete the measurement protocol. All participants also confirmed that they were not taking any medication, and refrained from drinking alcohol the day before measurement. Prior to participating in the study, each volunteer provided written informed consent, and the study was approved by the Institutional Review Board (19537-2).

### 2.2 Data collection

Each participant was asked to wear an elastic sportswear-type wearable (MATOUS^®^ VS, Teijin Frontier Sensing Ltd., Osaka, Japan) that is similar in comfort to exercise underwear. Prior to the experiment, the collegiate athletes confirmed that they felt no discomfort wearing it during sports activities. A data logger operating at 1000 Hz (SS-ECGHRAG, Sports Sensing Ltd, Fukuoka, Japan) was also attached to each participant’s upper back. An inertial sensor was built into the data logger (Figure 1), while two ECG electrodes were located on either side of the wearable. Both the ECG and acceleration signals were recorded by the data logger and transmitted to a laptop computer. Using a respiratory gas analyzer (AE-300S AEROMONITOR, Minato Medical Science Ltd., Tokyo, Japan), the breath-by-breath method was implemented to measure oxygen uptake 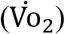, which is the gold standard physiological index. This allowed us to examine the feasibility of using the heart rate obtained by the sportswear-type wearable, as an indicator of exercise intensity. A linear relationship has been reported between heart rate and oxygen uptake [17, 18]; therefore, the use of oxygen uptake which is the gold standard for evaluating physiological intensity is appropriate for a feasibility study.

**Figure 1:**
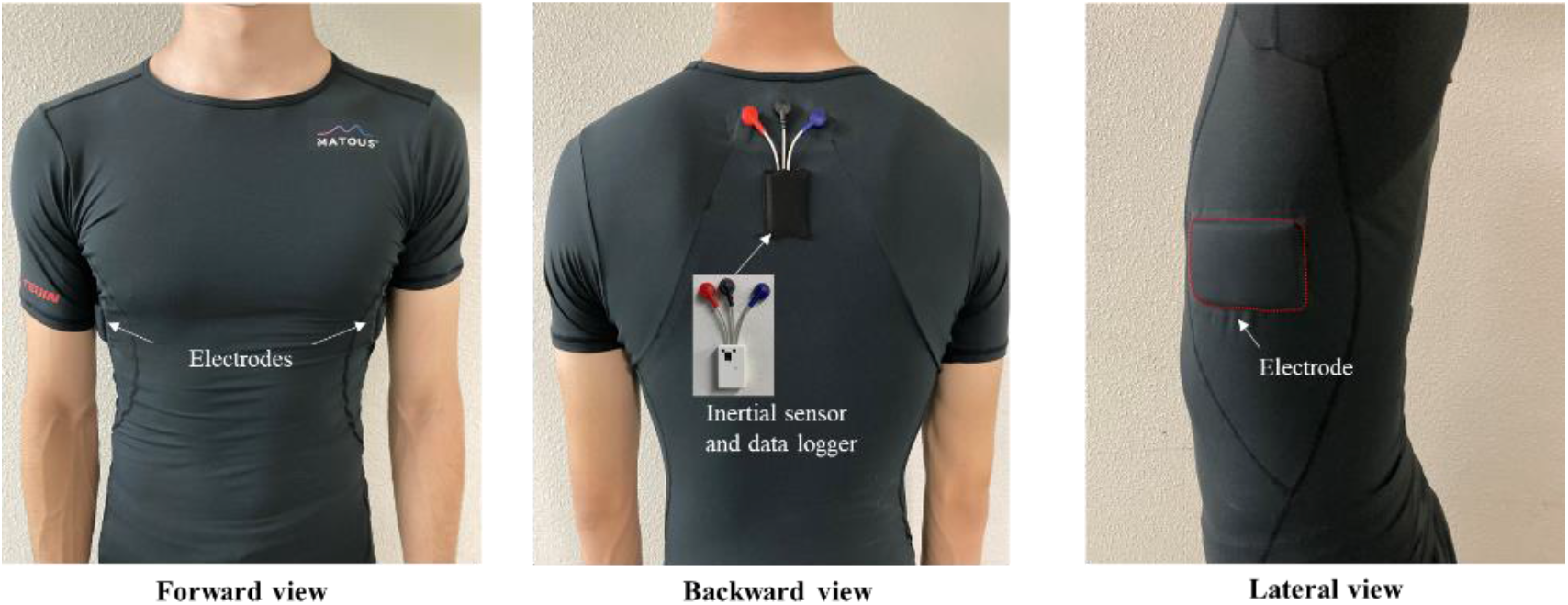
Sportswear-type wearable. ECG electrodes are located under each armpit, and the inertial sensor embedded into the data logger is mounted on the upper back.

A linear incremental loading test was performed using a treadmill (Elite T5000, Johnson Health Tech Japan Ltd., Osaka, Japan) set at a zero-degree incline. After recording the measurement at rest (standing), the participants were made to walk (1-6 km/h), jog (7-12 km/h), and run (13-20 km/h); speed was increased by 1 km/h every 30s. All measurements were taken in an indoor laboratory. This allowed us to examine the mechanical feasibility of evaluating the acceleration index under an exercise, intensity-controlled, condition.

### 2.3 Data analysis

The acceleration’s x-axis, y-axis, and z-axis were measured by the inertial sensor and then filtered by a Butterworth band-pass filter (0.5 Hz - 20 Hz), allowing the norm of acceleration 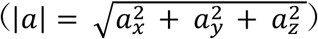 to be calculated. Then the norm of acceleration was filtered by a moving average filter with a 1-second window, defined as acceleration index. Following a Butterworth high-pass filtering process with a cutoff frequency of 5 Hz, the ECG signal was also filtered using a fourth order Savitzky–Golay filter. The filtered ECG data were used to detect the R-R interval and calculate the heart rate per minute (bpm). Then the calculated heart rate was filtered by a moving average filter with a 1-second window. Oxygen uptake was averaged every 30s using the respiratory gas analyzer software on the output of each breath, from rest to exercise completion. Data analysis was then performed using a custom-made MATLAB R2020a program (MathWorks, Inc.).

### 2.4 Statistical analysis

Heart rate, acceleration index, and oxygen uptake during the last 15s were averaged and used as representative values for each participant within each experimental condition. The mean values and standard deviations across all participants were determined using descriptive statistics. The relationship between heart rate and oxygen uptake was determined using a single regression analysis. To examine whether the acceleration index is equivalent to a change of 1 km/h on the treadmill or if the index adequately reflects the change in exercise intensity, single regression analyses between acceleration indices and treadmill speeds were performed for each participant, for the following exercise types: low-intensity (walking: 1-6 km/h); medium-intensity (jogging: 7-12 km/h); and high-intensity (running: 13-20 km/h). A one-sample t-test was then used to check whether the slope of the regression line was significantly greater than zero.

## 3 Results

The ECG signals obtained from the sportswear-type wearable were capable of detecting the R wave, necessary for calculating the heart rate over a wide range of exercise intensities: low (walking), moderate (jogging), and high (running) (Figure 2). The physiological indices (heart rate and oxygen uptake) showed linear trends with increasing treadmill speed. Similarly, the acceleration index showed linear trends for all exercise intensities (Figure 3). However, a rapid change was observed at the switching point between low-intensity exercise (walking) and moderate-intensity exercise (jogging) (Figure 3). The single regression analysis between heart rate and oxygen uptake showed that the slope of the regression line was significantly greater than zero (p < 0.001), with a high coefficient of determination (R^2^ = 0.96) (Table 1, Figure 4). Another single regression analysis between treadmill speed and the acceleration index showed that the slope of the regression line was significantly greater than zero at all intensities (p < 0.001) (Table 1, Figure 5), with a high coefficient of determination (walking, 0.95; jogging, 0.96; running, 0.90) (Table 1).

**Table 1:** Slope of regression lines and coefficient of determination

| | acceleration index | | | bpm and $\dot{V}O_2$ |
| --- | --- | --- | --- | --- |
|  | walking | jogging | running |  |
| slope of regression line | 0.06<br>(0.05-0.07) | 0.06<br>(0.04-0.08) | 0.02<br>(0.01-0.03) | 0.54<br>(0.47-0.62) |
| coefficient of determination<br>of regression line ($R^2$) | 0.95<br>(0.86-0.99) | 0.96<br>(0.93-0.99) | 0.90<br>(0.77-0.99) | 0.96<br>(0.90-0.99) |

**Figure 2:**
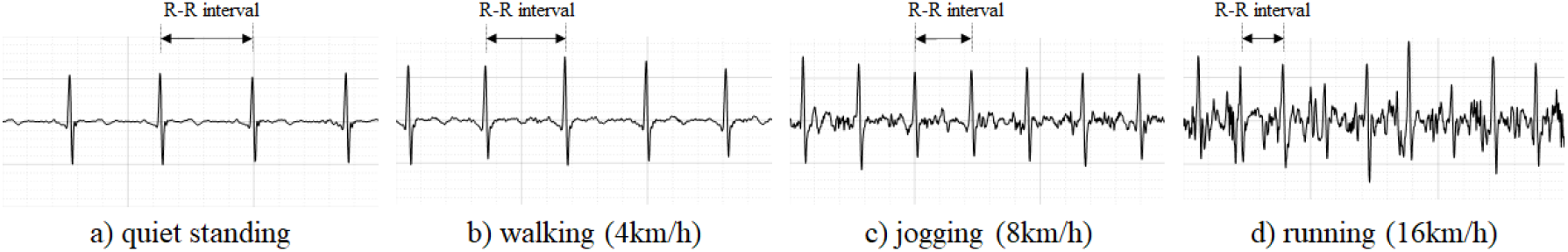
ECG waveforms recorded by the sportswear-type wearable: (a) at rest (quiet standing); (b) walking; (c) jogging; and (d) running.

**Figure 3:**
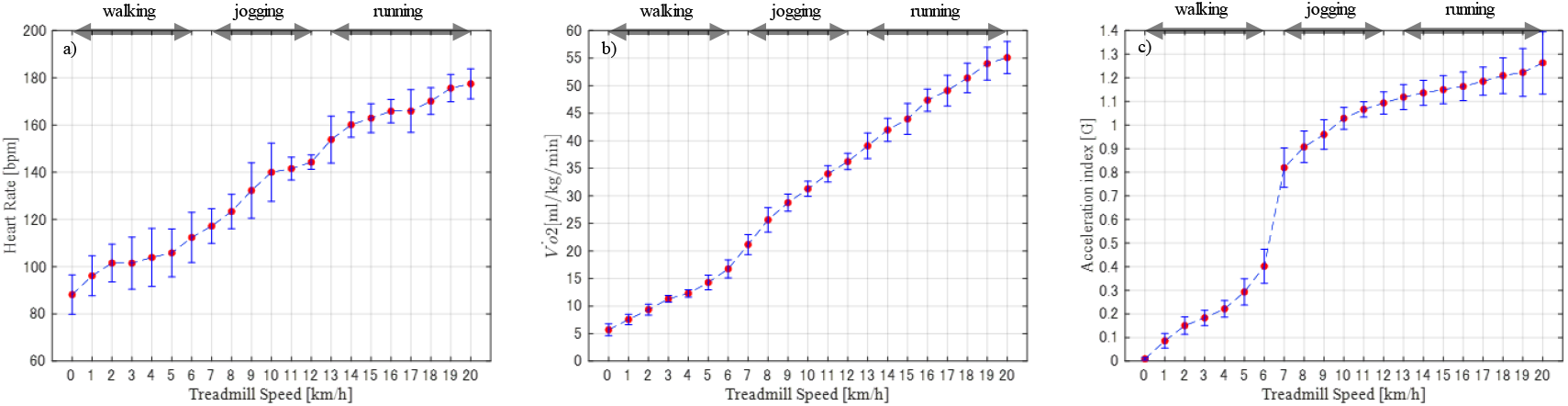
Mean and standard deviation of (a) heart rate (bpm), (b) oxygen uptake 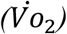 and (c) acceleration index at each treadmill speed across all participants.

**Figure 4:**
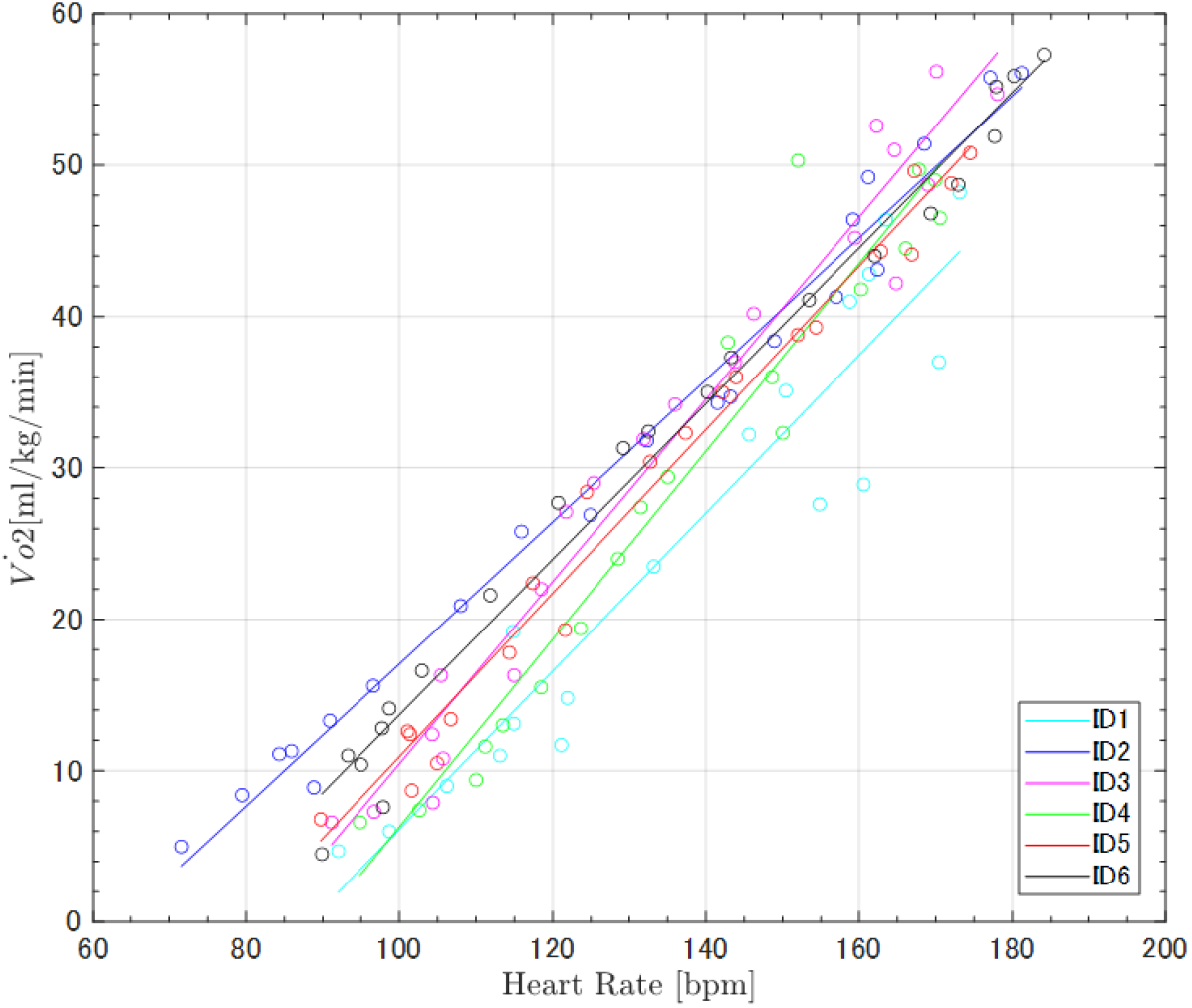
Regression lines of the heart rate and oxygen uptake comparison of all participants. All participants showed a linear relationship with small variations (r ≥ 0.96).

**Figure 5:**
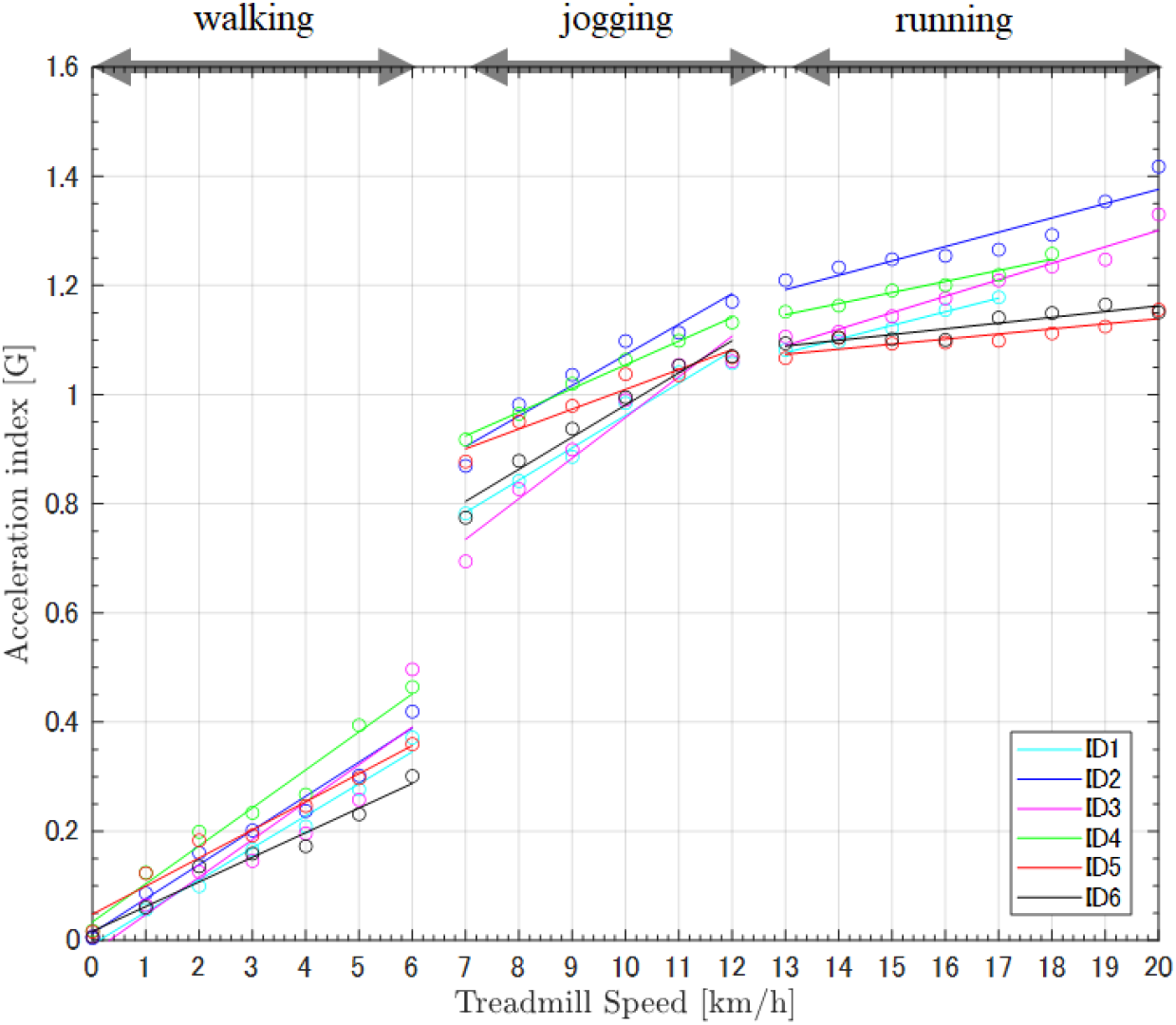
Regression lines of the acceleration index and treadmill speed comparison of all participants. All participants showed linear trends between acceleration index and treadmill speed within each condition: walking (0-6 km/h); jogging (7-12 km/h); and running (13-20 km/h).

## 4. Discussion

We examined the feasibility of evaluating exercise intensity across wide range of physical activity (quiet standing, walking, jogging, and running), using a sportswear-type wearable in a controlled laboratory setting. The results of this study demonstrated that (1) the heart rate recorded by the sportswear-type wearable highly correlated with the oxygen uptake and (2) the acceleration index systematically increased alongside the rise in exercise intensity. We suggest that it is feasible to use sportswear-type wearables to evaluate exercise intensity across different physical activities.

The sportswear-type wearable allows the detection of R-R intervals on the recorded ECG waveforms during quiet standing, walking, jogging, and running (Figure 2). This suggests that heart rate can be determined from the sportswear-type wearable, and bpm is an appropriate physiological indicator of exercise intensity across activities. The signal-to-noise ratio decreases with the increase in artifacts during moderate to high intensity movements, such as jogging and running (Figure 2). The artifacts may be a result of the wearable’s motion artifact and contaminated muscle activity. Recently, wearable sensors have been used to predict and detect cardiac diseases [26]. With the current accuracy of ECG measurements by the sportswear-type wearable, it is not suitable to detect arrhythmia during high intensity exercise. However, it is assumed that it may be possible at rest and low intensity exercise. In future studies, it is necessary to develop sportswear-type wearable that can detect arrhythmia during high intensity exercise and sports activities.

ECG-derived heart rate demonstrated a positive trend with increasing exercise intensity (Figure 3 and 4). Although the validity of the ECG recording could not be evaluated by comparing its results with other ECG recording devices, the strong association between heart rate and oxygen uptake identified in this experiment indicates that the heart rate obtained from the sportswear-type wearable is adequate for evaluating physiological intensity. Indeed, prior studies have also reported a linear relationship between heart rate and oxygen uptake [27, 28], reinforcing the validity of the heart rate recorded by our sportswear-type wearable. Validation studies on the heart rate determined via optical watch-type wearable devices have reported low accuracy during activity with varying intensity [29]. This suggests that it is difficult to use watch-type wearables in competitive sports that involve intense activity. Currently, the sportswear-type wearables demonstrate an acceptable level of accuracy for detecting R-R intervals, indicating that the calculated heart rate has corresponding accuracy. The sportswear-type wearable could thus be applied to quantify physiological intensity during sports of wide-ranging intensities, from rest to high intensity.

The linear association between the acceleration index and controlled treadmill speeds (walking, jogging, and running) (Figure 3 and 5) demonstrates that the acceleration index recorded by the wearable’s inertial sensor is adequate for detecting changes in physical intensity. A previous study had reported a change in acceleration index when going up to a speed of 16.5 km/h; conversely, our experiment’s participants reached a greater speed (up to 20 km/h), which is common to many sports activities. In a previous study, the rapid change of acceleration index during the transition from waking to jogging was recorded by both the inertial sensor located on the upper back, and the wearable’s sensor on the lower back [30]. The regression line’s slope was smaller during the running condition than during walking and jogging. This demonstrates that the sensitivity of the acceleration index relative to the change in activity intensity may slightly decrease when the athlete reaches running and sprinting intensity. However, the slope of the regression line was significantly greater than zero; therefore, the results suggest that the acceleration index recorded by the inertial sensor located on the upper back is useful for the evaluation of varied intensity in sports activities. GPS-recorded moving velocity has been widely used as an indicator of exercise intensity during competitive sports, especially team sports [30-32]. However, GPS validation studies have reported that the technology’s reliability and accuracy significantly reduce when individuals move fast over short distances, regardless of sampling frequency [33-35]. In addition, GPS devices are relatively large and heavy when compared with the accelerometers. Therefore, the small and light accelerometer on the upper back not only demonstrates feasibility for detecting fast-moving activity over a short distance, but also causes less discomfort, making the wearable device adequate for competitive sports.

This study had two major limitations. First, we could not directly compare ECG results between those obtained from the sportswear-type wearable and the ground truth measure. Instead, we verified the feasibility of heart rate as an indicator of physiological intensity by external comparison with oxygen uptake. In future studies, direct comparisons of ECG records will provide results that better indicate the possibility for using sportswear-type wearables to detect abnormal ECGs derived from cardiovascular disease. Second, this study only tested a sportswear-type wearable designed for men. However, if a sportswear-type wearable designed for women adopts the same structural locations for the ECG electrodes and inertial sensor, we believe that a similar study to ours with female participants will also demonstrate feasibility for evaluating exercise intensity.

## 5. Conclusion

The major advantage of sportswear-type wearables is that it is as comfortable as sportswear, which enables athletes to wear them even in competitive sports. We demonstrated that a sportswear-type wearable, with two integrated ECG electrodes and an inertial sensor, is adequate for evaluating exercise intensity across physical activities that range from rest to high intensity. It suggests the sportswear-type wearable is thus suitable for the monitoring of physical and physiological exercise intensity during various exercise types and sports activities. The sportswear-type wearables will be used as assistive devices to ensure sport-related safety and improve athletic performance.

## Data Availability

All data produced in the present study are available upon reasonable request to the authors

## Author contribution

Conceptualization, Y.M., I.O., E.M., and K.N.; Data acquisition, Y.M.; Data analysis, Y.M, S.K., and I.O.; Interpretation, Y.M., I.O., K.Y., T.Y., E.M., and K.N. Writing original draft, Y.M., S.K. I.O. All authors critically revised the report, commented on drafts of the manuscript, and approved the final report.

## Institutional Review Board Statement

The study was conducted in accordance with the guidelines of the Declaration of Helsinki and approved by the Institutional Ethics Committee of Osaka University Hospital (19537-2, July 1, 2020).

## Informed Consent Statement

All volunteers provided written informed consent.

## Data Availability Statement

The data analyzed in this manuscript will be made available from the corresponding author upon reasonable request.

## Acknowledgements

Not applicable

